# Beyond Benchmarks: Towards Robust Artificial Intelligence Bone Segmentation in Socio-Technical Systems

**DOI:** 10.1101/2025.06.11.25329022

**Authors:** Kunpeng Xie, Lennart Johannes Gruber, Martin Crampen, Yao Li, André Ferreira, Elias Tappeiner, Maxime Gillot, Jan Schepers, Jiangchang Xu, Tobias Pankert, Michel Beyer, Negar Shahamiri, Reinier ten Brink, Gauthier Dot, Charlotte Weschke, Niels van Nistelrooij, Pieter-Jan Verhelst, Yan Guo, Zhibin Xu, Jonas Bienzeisler, Ashkan Rashad, Tabea Flügge, Ross Cotton, Shankeeth Vinayahalingam, Robert Ilesan, Stefan Raith, Dennis Madsen, Constantin Seibold, Tong Xi, Stefaan Bergé, Sven Nebelung, Oldřich Kodym, Osku Sundqvist, Florian Thieringer, Hans Lamecker, Antoine Coppens, Thomas Potrusil, Joep Kraeima, Max Witjes, Guomin Wu, Xiaojun Chen, Adriaan Lambrechts, Lucia H Soares Cevidanes, Stefan Zachow, Alexander Hermans, Daniel Truhn, Victor Alves, Jan Egger, Rainer Röhrig, Frank Hölzle, Behrus Puladi

## Abstract

Despite the advances in automated medical image segmentation, AI models still underperform in various clinical settings, challenging real-world integration. In this multicenter evaluation, we analyzed 20 state-of-the-art mandibular segmentation models across 19,218 segmentations of 1,000 clinically resampled CT/CBCT scans. We show that segmentation accuracy varies by up to 25% depending on socio-technical factors such as voxel size, bone orientation, and patient conditions such as osteosynthesis or pathology. Higher sharpness, isotropic smaller voxels, and neutral orientation significantly improved results, while metallic osteosynthesis and anatomical complexity led to significant degradation. Our findings challenge the common view of AI models as “plug-and-play” tools and suggest evidence-based optimization recommendations for both clinicians and developers. This will in turn boost the integration of AI segmentation tools in routine healthcare.

## Introduction

With the ongoing digital transformation of healthcare, segmentation-based acquisition of anatomical and pathological structures has become an essential step in both clinical practice and research. Applications scenarios span over a wide field including diagnostic, image-guided radiotherapy and virtual surgical planning^1–3^. However, manual segmentation is still labor intensive and time-consuming. To address this issue, a large number of automatic segmentation methods for different structures have emerged in the last decades, and among them artificial intelligence (AI) models utilizing deep learning methods are the most promising ones^4–6^. In the segmentation of mandible for example, AI models have progressed beyond research settings and have begun to translate to clinical use as certified medical software in clinical practice^7–10^. However, despite decades of algorithmic advancements, there remains no standardized clinical integration protocol for AI segmentation models, leaving clinical integration a major challenge^5,11,12^.

This may be due to the technocentric paradigm that has been in place for decades of comparing and developing algorithms in different challenges to push the limits of performance and ultimately surpass human capabilities^13^. While this technocentric perspective has brought us powerful models and refreshed leaderboards, it often overlooks the complex socio-technical systems in which AI models are applied. In terms of clinicians, recent work shows that their adoption of AI generated results hinge on transparency, robustness, and real-world applicability—not benchmark metrics alone^14,15^. Additionally, in real-world situations, medical imaging data is often acquired prospectively based on specific clinical needs, including a wide range of possible imaging protocols as well as different patient factors. In this respect, a shift in perspective from a techno-centric preoccupation to a socio-technical perspective^16^, which explicitly considers clinical contexts such as diverse imaging protocols, patient demographics, and practical workflow integration, would be highly beneficial in facilitating the effective translation of AI segmentation models into clinical routines and research settings.

Consequently, we need to understand how socio-technical factors affect the performance of AI segmentation models in general. A previous study found that factors such as the imaging modalities (e.g., CT and CBCT), scanning devices, and the reconstruction protocols (e.g., voxel size, thickness, convolutional kernels) all may impact segmentation outcomes^17^. While some studies have begun to explore these factors, previous studies have either focused on limited factors or used only a single AI model, leaving a comprehensive understanding of these interactions largely unveiled^18,19^.

To address this issue, instead of simply comparing models’ performance, we evaluated the impact of socio-technical factors on the overall performance of multiple AI models in this study. For this purpose, we chose the mandible, which is morphologically complex and a representative in bone segmentation, as the segmentation target and created a benchmark dataset that balanced both patient and imaging features. Notably, our study recruited the largest number of AI models for mandible segmentation evaluated to date. By systematically resampling the original data, we could experimentally control the impact of different factors as they would be controllable during medical image acquisition. We then evaluated the segmentation results to explore the general impact of imaging, patient, and anatomical region factors on the model performance. Based on the results, we further suggest best practice recommendations for clinicians in applying AI segmentation models. In addition, we put forward requirements for AI developers, who are expected to create next-generation models that are informed by the clinical challenges encountered in AI models. Our study provides a reliable evidence base for future clinical integration guidelines of AI segmentation models, helping bridge the gap between technical performance and practical deployment.

## Methods

In this multicenter study we evaluated state-of-the-art AI models from 20 different centers and companies around the world (Table 1). The study protocol was registered prospectively in the German clinical trial registry under registration ID DRKS00032736. All technical details can be found in this study protocol. The ethics application of the study was approved by the ethics committee at RWTH Aachen University (No.23-272). No informed consent was needed due to the use of anonymized retrospective patient data.

**Table 1.**
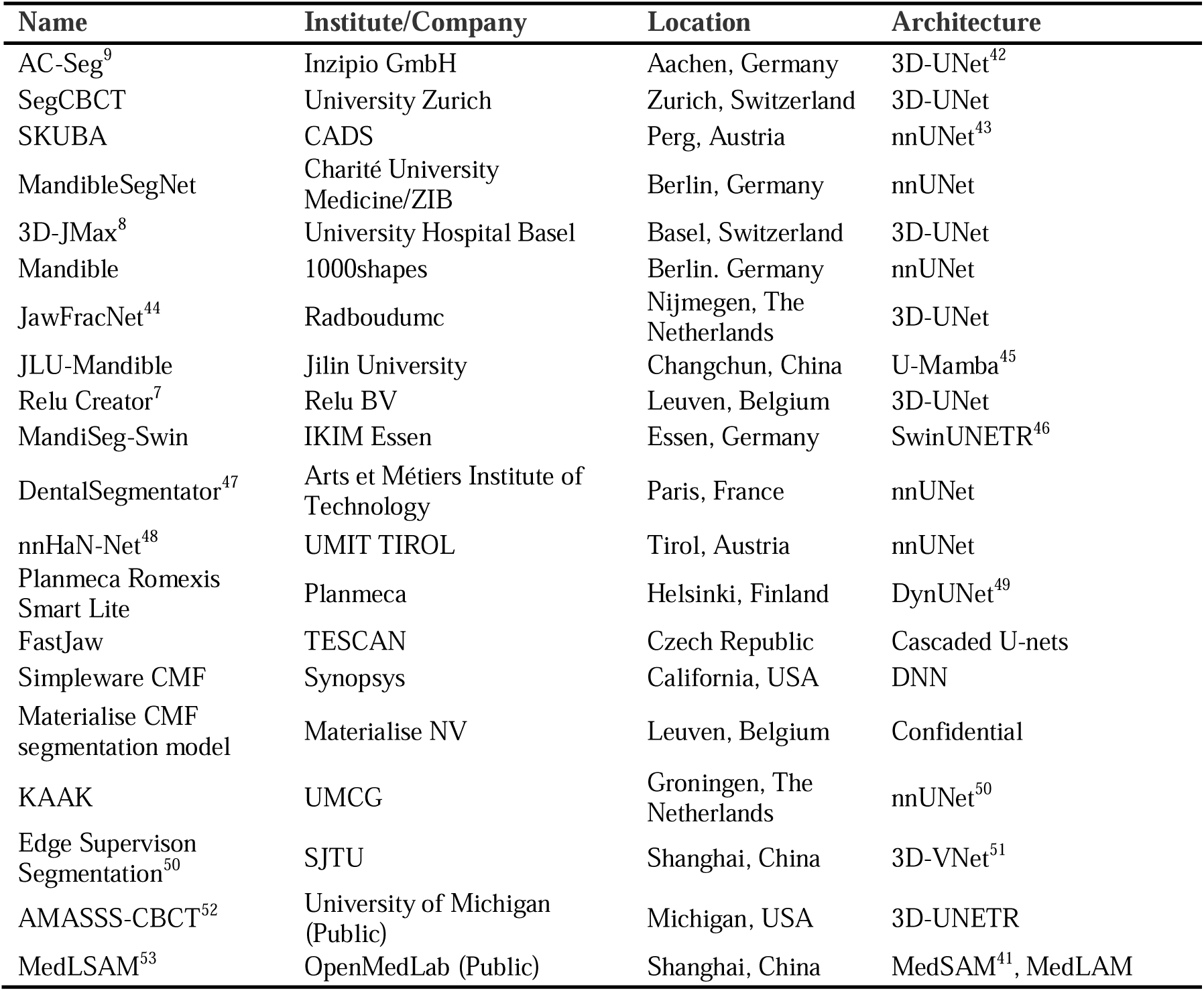
Summary of the recruited AI models.

### Dataset Preparation

To build a balanced benchmark dataset in terms of patient-related features, we selected 50 CT and 50 CBCT scans from 100 patients from a single center. In terms of patient characteristics, the sex ratio is 1:1 and the average age of patients was 48.47 years (range 19 – 91 years) (Supplementary Table 1). All selected scans were de-identified by cropping out the region above the inferior border of the orbital rim. Cases were excluded if cropping was not possible without affecting the condyle region. We systematically resampled the original 100 selected cases to create an additional 900 volumes, for a total of 1,000 volumes. This method, instead of selecting 1,000 cases directly, gave us full control over the voxel size, slice thickness, sharpness, noise and rotation of the mandible.

To obtain a balanced dataset, the features of the original CT/CBCT volumes were profiled prior to resampling. These features were quantified and measured in five aspects: a) voxel size (XY); b) slice thickness; c) sharpness; d) noise; e) rotation of the mandible. Where a) and b) were extracted from DICOM tags, c) was quantified via a Sobel-based edge intensity, and d) was derived from the standard deviation of the median-filter difference. Mandible rotations e) were calculated using bone landmarks. Based on the measurements, we chose five types of resampling methods namely: a) increase the slice thickness; b) expand the voxel size (XY); c) sharpening / smoothing; d) Gaussian-noise / denoise; e) rotation in axial, coronal and sagittal plane. A set of factors were tested and used in resampling these features respectively (Supplementary Table 4). By adjusting these factors, we have managed to approximate the distributions of features on the resampled dataset to the reference distribution from public datasets ^20–22^ or normal distribution. A total of 3,727,360 resampling combinations of imaging features were generated, from which 900 were randomly selected and resampled volumes were generated accordingly (Supplementary Figure 1). These down-sampled volumes, together with the initial 100 scans, resulted in a balanced final dataset of 1,000 volumes. The final distribution of patient and imaging features can be found in Supplementary Figure 1.

### Ground Truths

Mandible segmentations of the original scans were performed by two surgeons experienced in segmentation (KX and LG) independently in different software, KX in Mimics (Version 21.0) and LG in 3D Slicer (Version 5.6.2). The quality of segmentations was checked and approved by a third surgeon (BP). The principle of the segmentation was to preserve the anatomical bone structure of the mandible. In this case, all teeth, including dental implants, crowns and bridges, were segmented along with the mandible. Osteosynthesis materials (e.g. reconstruction plates, fixation screws/plates) were excluded in the segmentation, except for the part inside the mandible. The cancellous bone and the mandibular nerve canal were filled in so that the final segmentation result is free of internal cavities. Since resampling is not changing the anatomy of the bone, we applied the same resampling protocol in voxel scaling and rotation to the original ground truths to obtain corresponding segmentation results for the resampled 900 cases.

### Model Recruitment

The segmentation models included in this study need to meet the following criteria: a) deep learning based fully automatic segmentation tool; b) developed within the last five years; c) the output of the model is the mesh model or label map of the whole mandible; d) already trained and ready to use. Based on the literature study of a systematic review, we listed a group of models available in publications and searched further in online databases for other models published after the systematic review^5^. We contacted 35 corresponding authors and ten of them agreed to participate in the study. In addition, ten companies that offer mandible segmentation tools as a service were contacted. Eight of them joined our study. Furthermore, we have searched public repositories for available models and applied two trained models. With a data transfer agreement (DTA), the final dataset was shared with the collaborators, and segmentation results were returned to RWTH Aachen for evaluation. If a DTA was not feasible or the model was publicly available, inference was conducted locally at RWTH Aachen University.

### Evaluation

To further evaluate the segmentation quality in different anatomical regions of the mandible, we delineated nine ROIs by K.X and controlled by B.P.: condyle L/R, inferior alveolar nerve (IAN) entrance L/R, IAN exits L/R, dentition, inferior border. The last ROI, mandible body, was defined as the rest of mandible excluding the ROIs. All of the above ROIs were created based on reference points manually labelled on the volume by KX. Segmentation results were compared to both manual ground truths and the mean value was taken as the final result. We chose four metrics for evaluation: DSC, NSD, HD95, and MASD, and all metrics were calculated using the python package from Nikolov et al.^23,24^ on the whole mandible and on all ROIs respectively. No evaluations in the dentition region were conducted if the AI model cannot segment the teeth. All evaluations were conducted anonymously to secure the interests of all researchers and companies.

### Statistical analysis

The statistical analysis was conducted with the R programming language (Version 4.4.2). For descriptive statistics on data with non-normal distribution, we applied the non-parametric Mann-Whitney U test to evaluate statistical significance, followed by a bootstrap procedure with 5000 replicates to obtain the 95% CI for the median difference. Factors listed in the above section were set as fixed effect in the LMM while the difference of the AI models was considered as random effect. We checked the collinearity of selected fixed effects and found that sharpness and noise were highly corelated with a Variable Inflation Factor (VIF) of 10.943. In this case, noise was removed from the list of factors. We scaled the factors and tested multiple combinations of settings and selected one optimal LMM for each ROI and the whole mandible on each metric. LMM results on DSC are displayed in Figure 7. LMMs on other metrics and details of fitted models are described in Supplementary Figure 2, 3 and 4. We performed further analyses of the models described above to establish the evidence base for our recommendations.

## Results

### Recruited AI Models and overall segmentation results

A total of 20 commercial and research AI models for mandible segmentation from different countries across the world were recruited in this study, with the workflow shown in Figure 1. All models were developed over the last 5 years and listed in Table 1. Due to privacy reasons of the participating companies, evaluations of these models were anonymized. The evaluation was performed on ground truths of two investigators with an interrater correlation of 95.7% in Dice Similarity Coefficient (DSC, i.e. overlap measurement). From the 1000 volumes to be segmented, on average 942 volumes were successfully segmented and 19,218 segmentations were evaluated. The model designations are listed in descending order according to the number of volumes with DSC greater than 90% in their segmentation results (Fig. 2a). Only one model (S) was unable to segment any CBCT volume.

**Figure 1.**
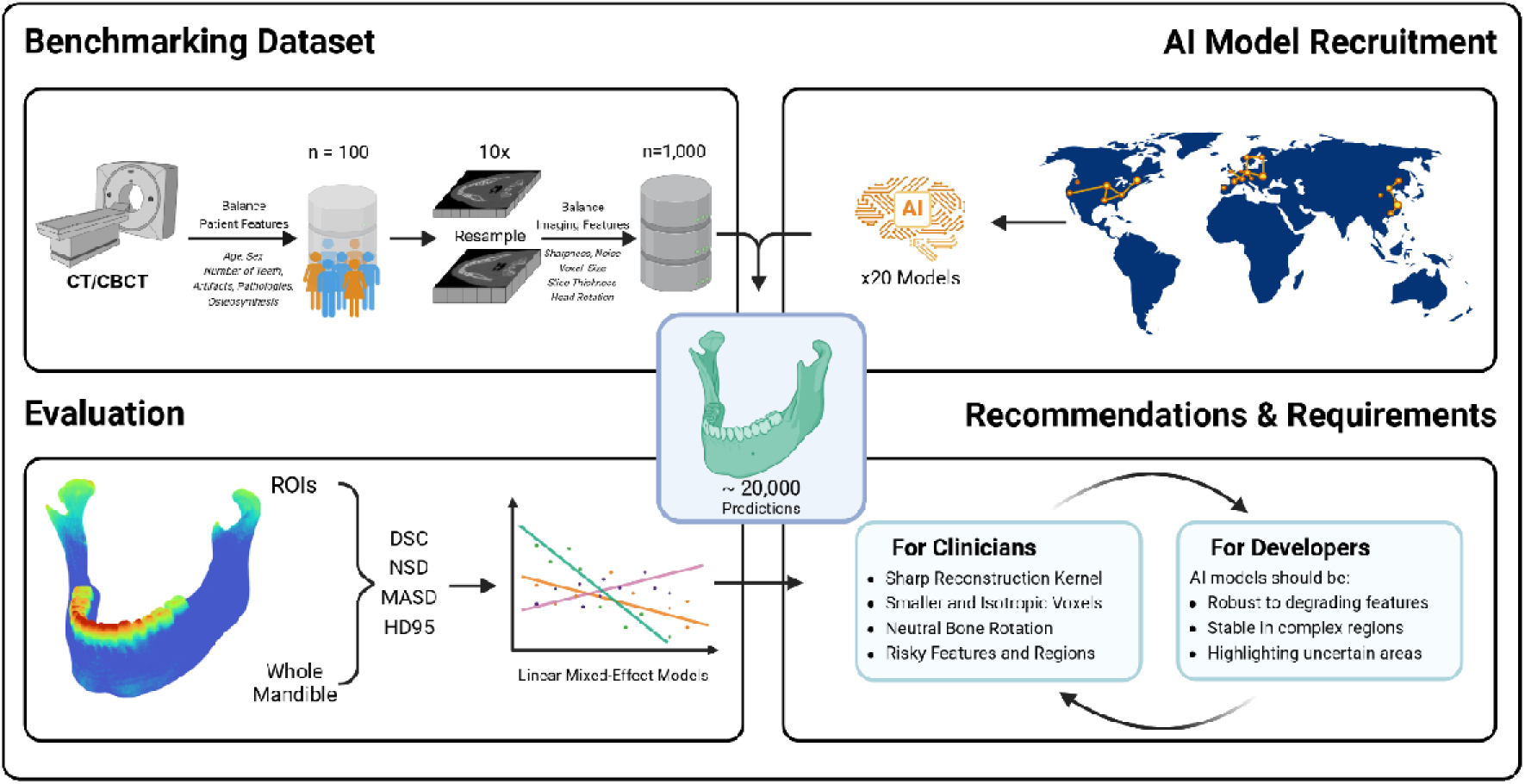
Workflow of the study. Created with BioRender.

**Figure 2.**
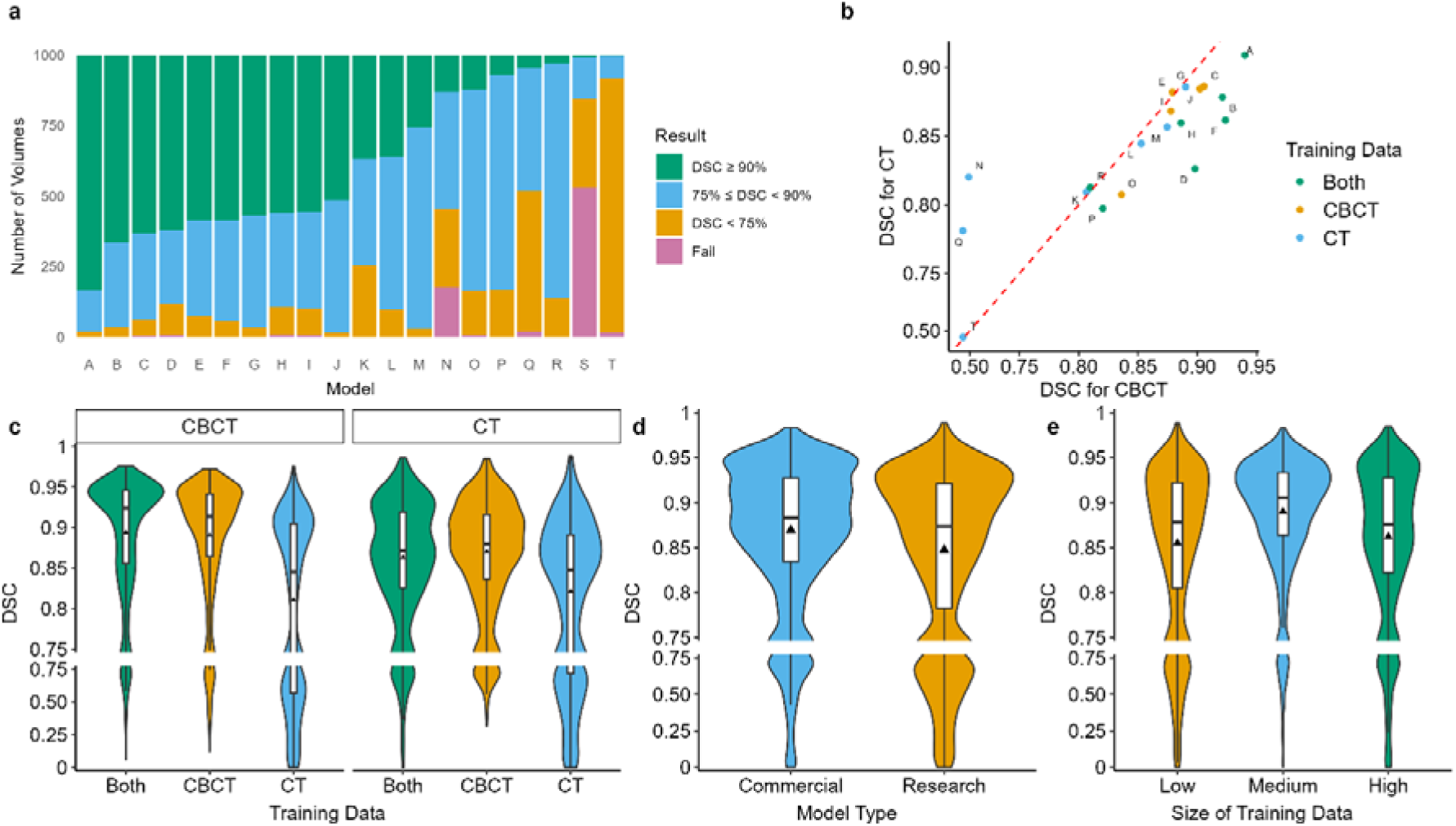
Model related factors and segmentation performance. (**a**) Ranking of models based on segmentation quality. Decrease by number of good cases (DSC ≥ 0.9) (**b**) Distribution of model performance in CT and CBCT subsets based on mean DSC (**c**) Impact of training data on overall segmentation performance (**d**) Impact of model type (**e**) Impact of the size of training dataset. Low: 0-150 cases; Medium: 150-300 cases; High: 300+ cases.

Table 2 presents the overall performance of the models, including the CT and CBCT subsets. The metrics used were: DSC as primary metric, Normalized Surface Dice (NSD, i.e. boundary agreement), 95 percentile Hausdorff Distance (HD95, i.e. worst-case boundary error), and Mean Average Surface Distance (MASD, i.e. average boundary deviation)^25^. The mean values of DSC and NSD for all models are both 81.7%. While the mean values of HD95 and MASD are 14.89 mm and 2.73 mm, respectively. Model *A* demonstrates the best performance across almost all metrics. We explored the effect of the type of training data on the segmentation results (Fig. 2b, c). It is interesting to note that the models trained with only CBCT data show better results than the models trained with only CT data (Mann-Whitney U test, p < 0.001), and the median difference was estimated as 5.10% with a 95% bootstrap confidence interval (CI) of [4.71%, 5.51%]. Yet the difference is not significant between CBCT and combination of both data modalities (Mann-Whitney U test, p = 0.733). Commercial models demonstrate better performance compared to research models (Mann-Whitney U test, p < 0.001), with a median difference of 1.03% [95% CI: 0.75%, 1.34%]. Regarding the amount of training data, the models trained on a moderate number of scans (150–300 cases) exhibited the optimal segmentation performance among all groups (p < 0.001, Kruskal-Wallis test; Fig. 2d,e). The median DSC difference between the medium and low groups was 2.70% [95% CI: 2.39%, 2.97%], and between the medium and high groups was 2.87% [95% CI: 2.55%, 3.16%].

**Table 2.** Segmentation performance (mean ± sd) of AI models on the whole mandible. Best performances were marked in blue. Model S failed to segment CBCT volumes. Models anonymized by descending order of number of segmentations with DSC > 90%.

| Model | DSC (%) |  |  | NSD (%) |  |  | HD 95 (mm) |  |  | MASD (mm) |  |  |
| --- | --- | --- | --- | --- | --- | --- | --- | --- | --- | --- | --- | --- |
|  | CT | CBCT | Overall | CT | CBCT | Overall | CT | CBCT | Overall | CT | CBCT | Overall |
| A | 90.87±<br>9.19 | 94.01±<br>4.86 | 92.44±<br>7.51 | 93.77±<br>9.80 | 94.37±<br>5.55 | 94.07±<br>7.97 | 4.05±<br>20.70 | 2.26±<br>5.05 | 3.16±<br>15.09 | 0.60±<br>3.02 | 0.29±<br>0.46 | 0.44±<br>2.16 |
| B | 87.82±<br>7.03 | 92.11±<br>5.48 | 89.97±<br>6.66 | 89.56±<br>9.84 | 91.93±<br>7.02 | 90.75±<br>8.62 | 4.71±<br>11.64 | 3.90±<br>10.74 | 4.31±<br>11.20 | 0.64±<br>1.17 | 0.60±<br>1.78 | 0.62±<br>1.50 |
| C | 88.60±<br>7.22 | 90.57±<br>8.87 | 89.59±<br>8.15 | 91.34±<br>9.60 | 91.90±<br>9.60 | 91.62±<br>9.60 | 7.29±<br>28.00 | 5.08±<br>12.31 | 6.18±<br>21.64 | 1.71±<br>11.48 | 0.72±<br>1.99 | 1.21±<br>8.25 |
| D | 82.62±<br>18.74 | 89.81±<br>11.81 | 86.24±<br>16.03 | 83.40±<br>20.21 | 89.50±<br>12.03 | 86.48±<br>16.87 | 19.90±<br>56.52 | 4.68±<br>9.96 | 12.23±<br>41.12 | 3.15±<br>11.08 | 0.70±<br>1.76 | 1.91±<br>7.99 |
| E | 88.19±<br>6.92 | 87.91±<br>12.35 | 88.05±<br>10.01 | 89.18±<br>9.48 | 85.71±<br>13.66 | 87.44±<br>11.88 | 10.23±<br>33.46 | 11.32±<br>23.69 | 10.77±<br>28.98 | 1.45±<br>5.52 | 1.77±<br>4.24 | 1.61±<br>4.92 |
| F | 86.16±<br>8.05 | 92.37±<br>6.80 | 89.27±<br>8.07 | 88.60±<br>10.81 | 93.62±<br>8.52 | 91.11±<br>10.04 | 5.98±<br>13.37 | 3.08±<br>9.73 | 4.53±<br>11.78 | 0.78±<br>1.82 | 0.48±<br>1.51 | 0.63±<br>1.68 |
| G | 88.57±<br>4.81 | 89.02±<br>10.82 | 88.79±<br>8.37 | 90.04±<br>7.00 | 87.57±<br>10.76 | 88.80±<br>9.16 | 6.70±<br>14.40 | 8.13±<br>11.86 | 7.41±<br>13.20 | 0.62±<br>0.95 | 0.95±<br>2.41 | 0.78±<br>1.84 |
| H | 85.94±<br>12.05 | 88.63±<br>9.26 | 87.29±<br>10.81 | 87.80±<br>13.99 | 87.76±<br>10.85 | 87.78±<br>12.50 | 12.65±<br>29.67 | 9.47±<br>17.75 | 11.06±<br>24.49 | 2.81±<br>11.15 | 1.15±<br>2.27 | 1.98±<br>8.09 |
| I | 86.81±<br>8.61 | 87.77±<br>13.56 | 87.29±<br>11.35 | 88.66±<br>12.07 | 88.65±<br>15.19 | 88.65±<br>13.70 | 4.68±<br>11.40 | 9.26±<br>18.64 | 6.96±<br>15.59 | 0.61±<br>0.91 | 1.32±<br>3.11 | 0.96±<br>2.31 |
| J | 88.43±<br>3.98 | 90.22±<br>4.61 | 89.32±<br>4.39 | 90.45±<br>6.78 | 89.95±<br>6.30 | 90.20±<br>6.55 | 6.92±<br>22.98 | 5.92±<br>7.42 | 6.42±<br>17.07 | 0.74±<br>2.20 | 0.61±<br>1.04 | 0.68±<br>1.72 |
| K | 80.92±<br>18.31 | 80.63±<br>13.45 | 80.78±<br>16.06 | 83.16±<br>18.25 | 80.64±<br>12.93 | 81.90±<br>15.86 | 16.74±<br>26.51 | 10.19±<br>9.70 | 13.47±<br>20.22 | 2.15±<br>4.49 | 1.33±<br>1.35 | 1.74±<br>3.34 |
| L | 84.45±<br>10.07 | 85.28±<br>13.85 | 84.86±<br>12.11 | 86.58±<br>12.82 | 83.54±<br>13.65 | 85.06±<br>13.32 | 9.17±<br>21.49 | 7.70±<br>13.45 | 8.44±<br>17.93 | 1.19±<br>2.62 | 1.20±<br>2.34 | 1.20±<br>2.48 |
| M | 85.64±<br>4.10 | 87.45±<br>7.56 | 86.55±<br>6.15 | 87.12±<br>10.02 | 86.13±<br>9.80 | 86.63±<br>9.92 | 3.87±<br>9.59 | 5.18±<br>12.65 | 4.52±<br>11.24 | 0.61±<br>0.69 | 1.02±<br>3.22 | 0.82±<br>2.34 |
| N | 82.02±<br>15.89 | 49.38±<br>34.36 | 69.13±<br>29.55 | 83.12±<br>16.03 | 47.03±<br>32.13 | 68.87±<br>29.56 | 17.48±<br>61.10 | 52.16±<br>37.07 | 33.29±<br>54.35 | 2.90±<br>13.85 | 16.60±<br>16.09 | 9.14±<br>16.40 |
| O | 80.74±<br>8.61 | 83.61±<br>9.50 | 82.17±<br>9.17 | 80.76±<br>12.62 | 82.72±<br>10.81 | 81.74±<br>11.79 | 37.07±<br>47.93 | 10.63±<br>16.60 | 23.88±<br>38.24 | 5.12±<br>7.57 | 1.41±<br>2.83 | 3.27±<br>6.01 |
| P | 79.72±<br>11.64 | 82.02±<br>10.83 | 80.87±<br>11.30 | 83.31±<br>10.90 | 78.50±<br>9.30 | 80.91±<br>10.41 | 6.48±<br>15.68 | 7.38±<br>15.00 | 6.93±<br>15.34 | 0.86±<br>1.95 | 1.09±<br>2.49 | 0.98±<br>2.24 |
| Q | 78.11±<br>16.30 | 46.39±<br>27.64 | 62.59±<br>27.58 | 79.74±<br>15.64 | 47.31±<br>24.90 | 63.87±<br>26.28 | 15.75±<br>26.39 | 32.67±<br>24.16 | 24.05±<br>26.69 | 2.09±<br>4.19 | 6.43±<br>6.69 | 4.22±<br>5.96 |
| R | 81.26±<br>8.63 | 80.97±<br>10.36 | 81.12±<br>9.53 | 84.55±<br>8.06 | 79.54±<br>8.05 | 82.05±<br>8.43 | 5.83±<br>16.56 | 4.53±<br>4.46 | 5.18±<br>12.14 | 0.75±<br>2.69 | 0.64±<br>0.66 | 0.69±<br>1.96 |
| S | 50.71±<br>30.98 | NA | 50.71±<br>30.98 | 50.78±<br>30.43 | NA | 50.78±<br>30.43 | 105.38±<br>130.33 | NA | 105.38±<br>130.33 | 21.47±<br>29.62 | NA | 21.47±<br>29.62 |
| T | 47.23±<br>25.31 | 46.52±<br>20.26 | 46.87±<br>22.87 | 36.49±<br>21.60 | 34.73±<br>13.85 | 35.60±<br>18.11 | 64.49±<br>23.82 | 34.85±<br>5.88 | 49.53±<br>22.76 | 13.58±<br>9.00 | 7.99±<br>4.13 | 10.76±<br>7.52 |
| Overall | 81.38±<br>17.57 | 81.97±<br>20.02 | 81.66±<br>18.80 | 82.57±<br>19.64 | 80.74±<br>20.96 | 81.69±<br>20.31 | 17.97±<br>46.97 | 11.63±<br>19.88 | 14.89±<br>36.57 | 3.13±<br>10.34 | 2.31±<br>5.70 | 2.73±<br>8.42 |

### Imaging factors

Figure 3 shows the effect of imaging factors on segmentation performance of AI models. Higher sharpness level generally leads to better segmentation results (Fig. 3c). Further analysis in Linear Mixed-effect Models (LMMs) shows a 0.50% increase in DSC per 500 Hounsfield Unit (HU)/mm increase in sharpness (LMM, β = 0.001%, p < 0.001). However, the DSC improvements reached a plateau beyond a certain sharpness level (approximately 5000 HU/mm). This pattern was also observed regarding noise, where a moderate noise level led to the best segmentation performance. Larger voxel sizes in the XY plane significantly reduced segmentation performance, with a 0.16% decrease in DSC for every 0.1 mm increase in in-plane voxel size (LMM, β = –1.62%, p < 0.001). Increasing slice thickness also had a negative impact, with DSC declining by 0.10% for every 0.1 mm increase in slice thickness (LMM, β = –0.955%, p < 0.001). Rotation of the mandible in all three planes resulted in negative effects on segmentation performance, with axial and sagittal rotations reducing DSC by 0.51% and 0.69% per 5-degree increase, respectively (LMM, β*_axial_* = –0.102%, β*_sagittal_* = –0.138%, p < 0.001), while coronal rotation had no significance (p = 0.520).

**Figure 3.**
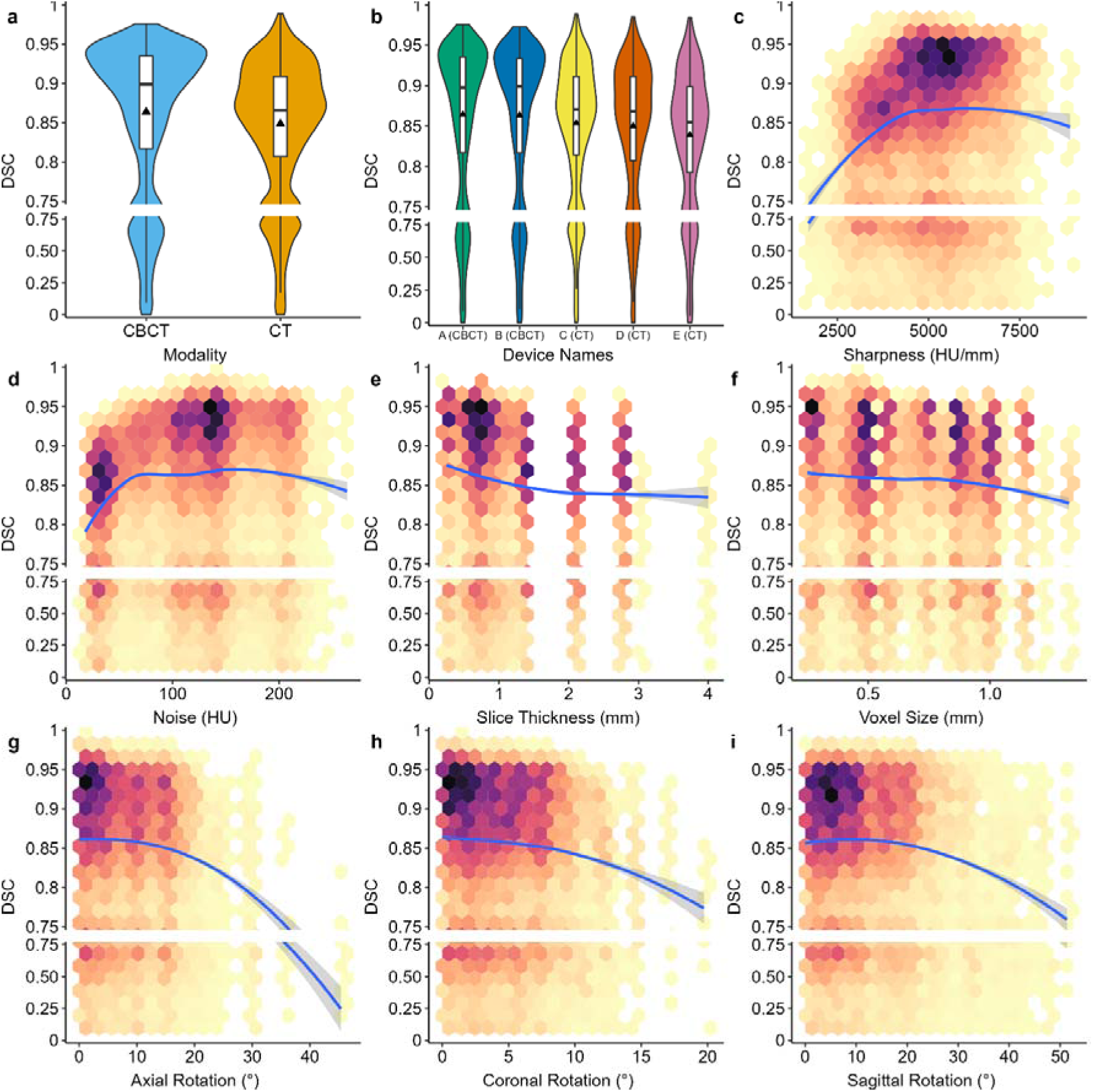
Image quality related factors and segmentation performance measured in DSC. (**a**) distribution of segmentation performance in CBCT and CT scans (**b**) segmentation performance in five devices used in the study (**c**) relationship between image sharpness and segmentation performance (**d**) the effect of image noise image noise and segmentation performance (**e**) relationship between slice thickness and segmentation performance (**f**) the impact of voxel size on segmentation performance (**g**) ∼ (**i**) the effect of bone rotation on segmentation performance. Colored hexagonal bins represent the distribution of data points. Darker colors indicate higher data density, while brighter colors indicate lower data density.

In univariable descriptive statistics, the AI models showed better performance on CBCT data than that of CT data (Mann-Whitney U test, p < 0.001; Fig. 3a), with a median DSC difference of 3.20% [95% CI: 2.96%, 3.45%]. For the use of different CBCT devices, no significant difference was found (Mann-Whitney U test, p = 0.198; Fig. 3b). Yet a marginal decline of 1.43% in median DSC [95% CI: 1.02%, 1.78%] is found in CT device C (Mann-Whitney U test, p < 0.001). However, in multivariable analysis the segmentation performance of the AI model on CT data is improved by 4.13% compared to CBCT data, (LMM, β = 4.129%, p < 0.001).

### Patient-related factors

Figure 4 displays the relationship between patient-related factors and segmentation performance. Male patients showed slightly better segmentation results than female patients, with a 1.0% higher DSC for males (LMM, β = 0.989%, p < 0.001). Older patients showed a decrease in DSC, but this effect was not significant (LMM, β = –0.011%, p = 0.126). We used the mean value of HU across the mandibular region to assess bone density and found that lower bone density reduced segmentation performance (Fig. 4c). The number of teeth in lower dentition positively influenced segmentation performance, with each additional tooth increasing DSC by 0.38% (LMM, β = 0.378%, p < 0.001). On the other hand, the presence of bone pathology (e.g. fractures, major cysts) reduced DSC by 0.71% (LMM, β = –0.708%, p < 0.05). Osteosynthesis material had the most significant negative effect, decreasing DSC by 7.90% (LMM, β = –7.90%, p < 0.001). Artifacts (e.g. metal, shadow) also negatively impacted segmentation, but showed no significant effect on DSC (LMM, β = –0.212%, p = 0.3313).

**Figure 4.**
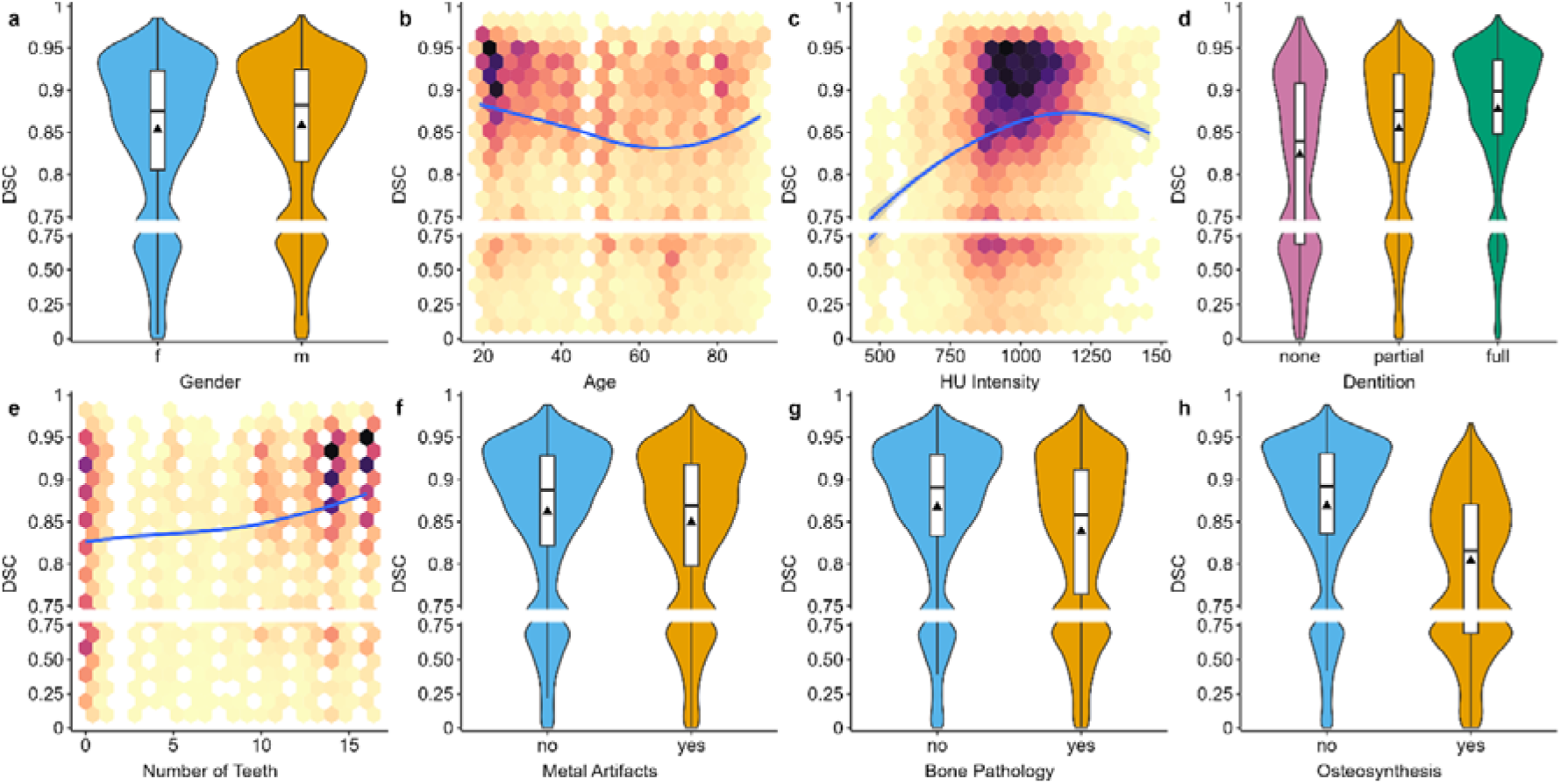
Patient related factors and segmentation performance. (**a**)Comparison of between female and male patients (**b**) relationship between age and segmentation performance (**c**)The effect of Hounsfield Unit (HU) intensity on segmentation performance (**d**)(**e**) The impact of dentition status and teeth count on segmentation performance **(f)** Comparison of segmentation performance between cases with and without metal artifacts **(g)** Influence of bone pathology on segmentation performance **(h)** The effect of osteosynthesis on segmentation performance

### Anatomical Regions

Figures 5 and 6 visualize the case-wise segmentation using heatmaps. Most errors can be observed in the condyle, dentition, and part of the mandibular body. The segmentation performance of the AI model is significantly degraded in regions of impaired mandibular continuity (Case 21,65), bone pathology (Case 16,61), and osteosynthesis material (Case 17,86) (Supplementary Table 3). The segmentation results in Table 3 further demonstrate the differences in segmentation performance across Regions Of Interest (ROIs). The mandibular body performed the worst in terms of HD 95 and MASD. In terms of DSC, the condyle in CBCT had the lowest score of 78.07%. In addition, the dentition also had the lowest NSD value of 84.16%, indicating a lack of accurate boundary segmentation in this region. In summary, the mandibular body has the highest segmentation error in the distance-based metrics, whereas the condylar and dentition regions exhibit the lowest DSC and NSD, respectively.

**Figure 5.**
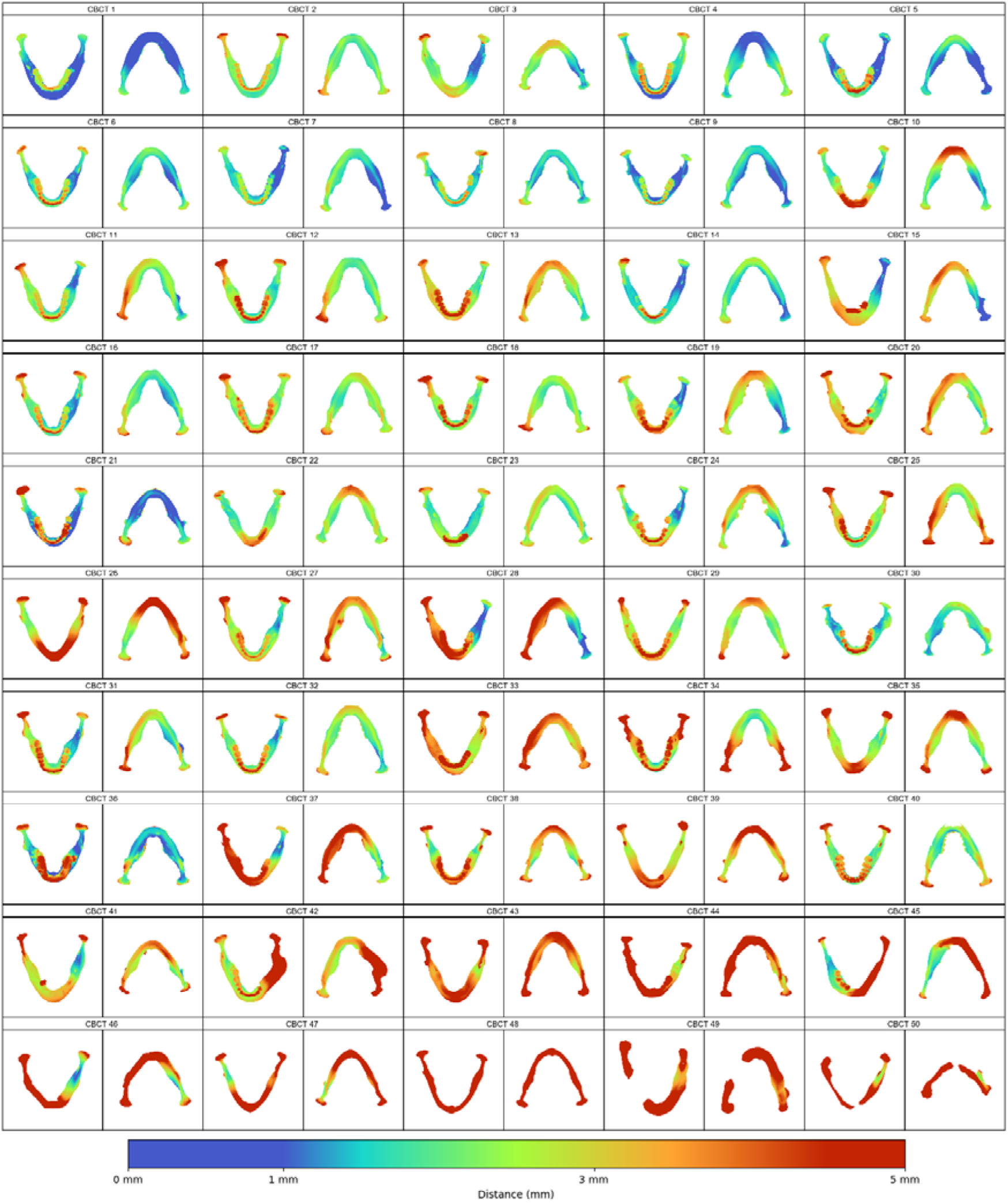
Heatmaps showing the average surface distance between AI segmentation results and the ground truths of CBCT scans. These segmentations were performed on the original scan and the 9 resample variants by 19 models (failed in model S), resulting in around 190 segmentations per case. Cases arranged in descending order of overall mean DSC.

**Figure 6.**
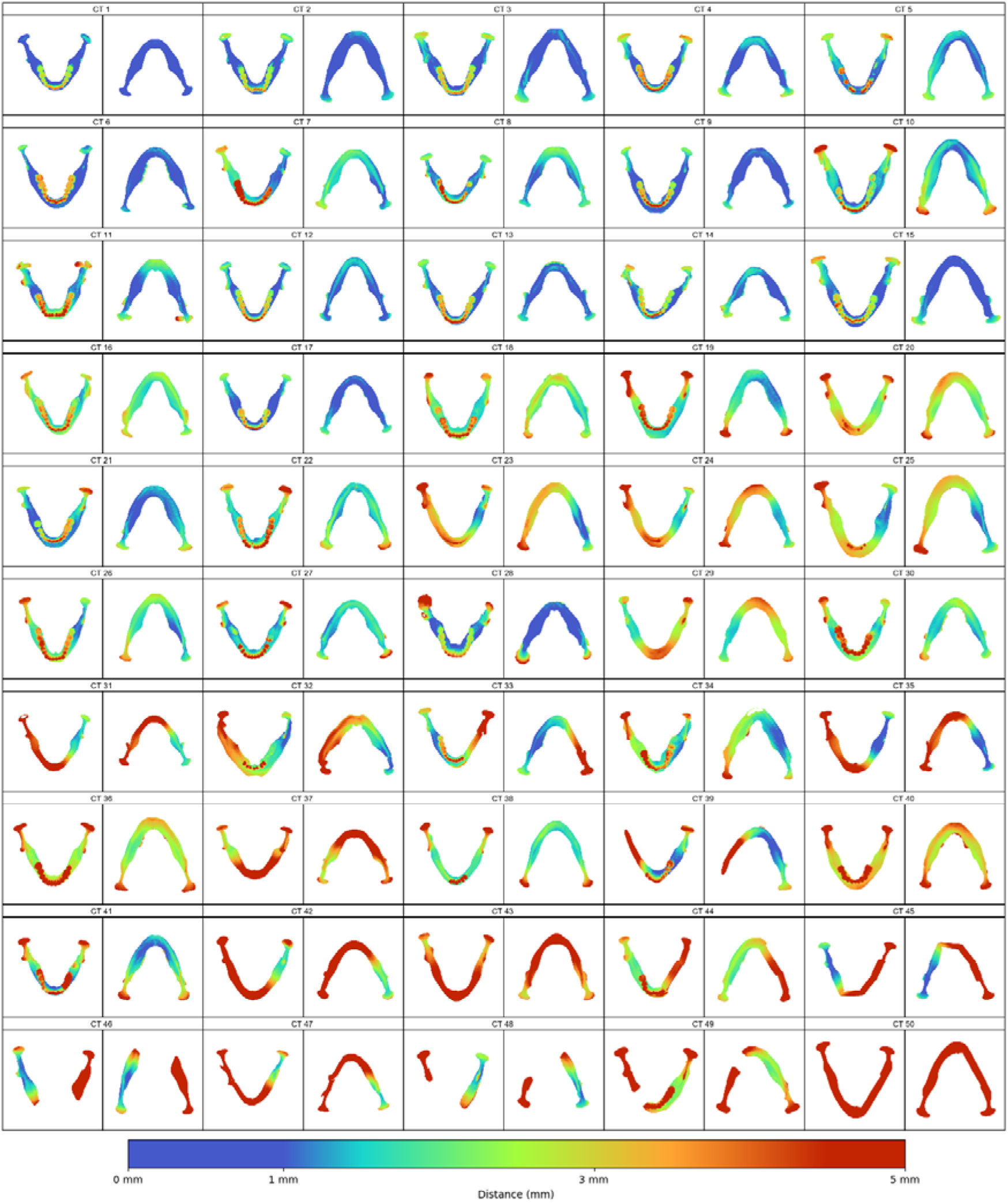
Heatmaps showing the average surface distance between AI segmentation results and the ground truths of CBCT scans. These segmentations were performed on the original scan and the 9 resample variants by 20 models, resulting in around 200 segmentations per case. Cases arranged in descending order of overall mean DSC.

**Figure 7.**
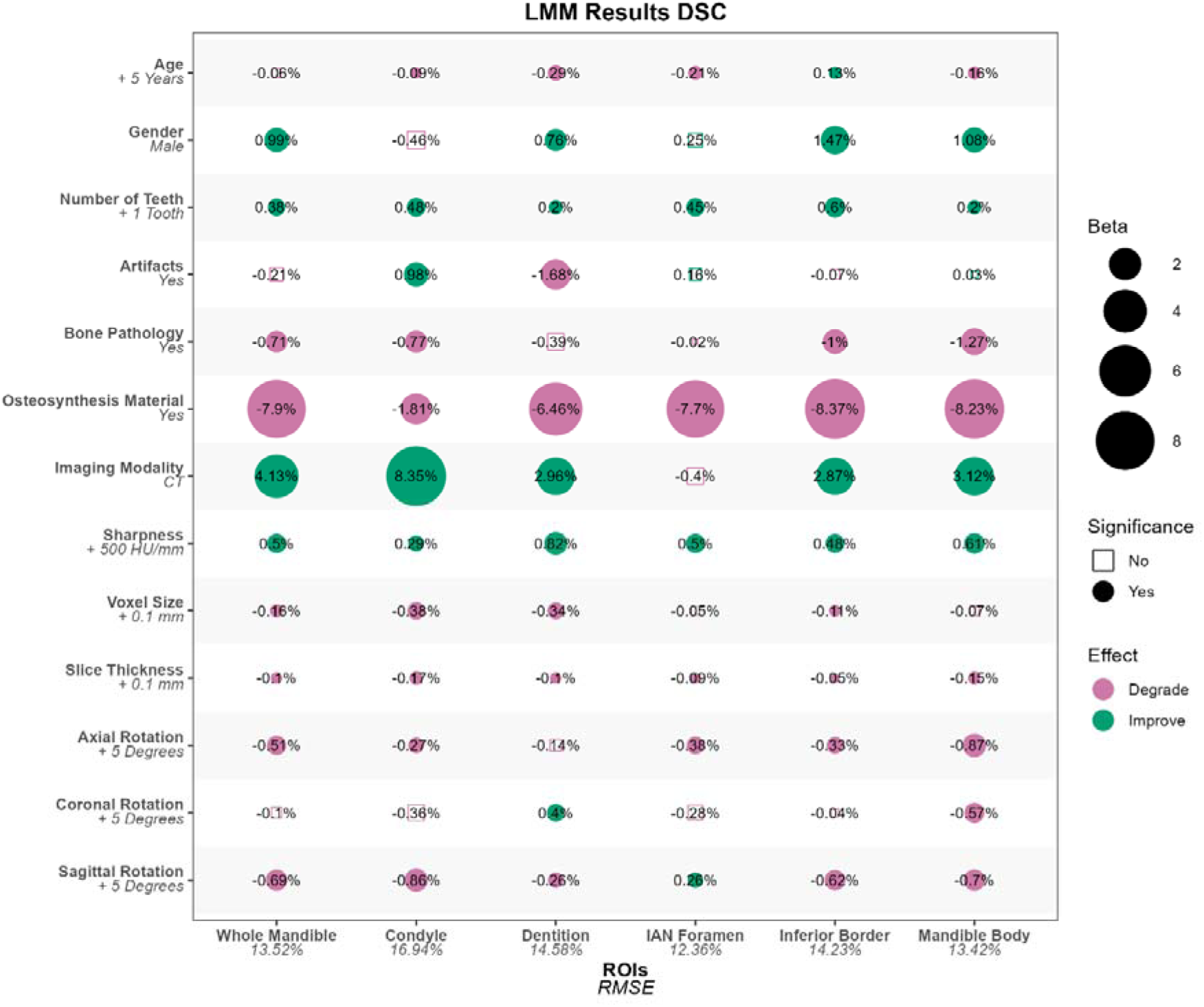
LMMs fitted on evaluation results in DSC% of five ROIs and the whole mandible. Factor considered significant when p<0.05.

**Table 3.** Performance of AI models (mean ± sd) on 5 anatomical regions and the whole mandible. Worst performances were marked in red.

| ROI | DSC (%) |  |  | NSD (%) |  |  | HD 95 (mm) |  |  | MASD (mm) |  |  |
| --- | --- | --- | --- | --- | --- | --- | --- | --- | --- | --- | --- | --- |
|  | CT | CBCT | Overall | CT | CBCT | Overall | CT | CBCT | Overall | CT | CBCT | Overall |
| Condyle | 82.26 ± 19.19 | 78.07 ± 23.54 | 80.26 ± 21.48 | 90.46 ± 17.37 | 82.24 ± 22.10 | 86.53 ± 20.19 | 2.24 ± 4.31 | 2.71 ± 2.92 | 2.47 ± 3.72 | 0.43 ± 0.89 | 0.62 ± 0.84 | 0.52 ± 0.87 |
| Dentition | 80.15 ± 19.31 | 83.13 ± 18.10 | 81.60 ± 18.79 | 85.41 ± 18.94 | 82.83 ± 18.58 | 84.16 ± 18.81 | 7.99 ± 26.86 | 5.42 ± 9.56 | 6.74 ± 20.41 | 1.51 ± 6.01 | 1.11 ± 2.81 | 1.32 ± 4.74 |
| IAN Foramen | 82.19 ± 14.45 | 85.58 ± 13.28 | 83.82 ± 14.00 | 95.94 ± 9.19 | 96.46 ± 8.77 | 96.19 ± 8.99 | 1.09 ± 1.02 | 0.93 ± 0.52 | 1.01 ± 0.82 | 0.12 ± 0.23 | 0.12 ± 0.15 | 0.12 ± 0.19 |
| Inferior Border | 84.64 ± 16.84 | 85.31 ± 18.81 | 84.97 ± 17.83 | 87.19 ± 17.44 | 84.08 ± 19.48 | 85.66 ± 18.53 | 10.11 ± 35.85 | 8.24 ± 15.90 | 9.19 ± 27.88 | 1.80 ± 7.74 | 1.54 ± 4.02 | 1.67 ± 6.19 |
| Mandible Body | 80.51 ± 18.36 | 82.37 ± 21.13 | 81.41 ± 19.78 | 85.02 ± 20.49 | 85.39 ± 21.93 | 85.20 ± 21.20 | 21.67 ± 60.00 | 10.23 ± 20.96 | 16.10 ± 45.76 | 4.30 ± 14.22 | 2.41 ± 6.67 | 3.38 ± 11.24 |
| Whole Mandible | 81.38 ± 17.57 | 81.97 ± 20.02 | 81.66 ± 18.80 | 82.57 ± 19.64 | 80.74 ± 20.96 | 81.69 ± 20.31 | 17.97 ± 46.97 | 11.63 ± 19.88 | 14.89 ± 36.57 | 3.13 ± 10.34 | 2.31 ± 5.70 | 2.73 ± 8.42 |

## Discussion

Although AI models have proven their performance, there are many open questions regarding the integration and limitations of current AI models in clinical routine as well as research. Recent qualitative research confirms that clinicians demand concrete insights into when and why AI fails in clinical settings, suggesting the need for comprehensive socio-technical evaluations^26^. Based on an experimental study with 20 current state-of-the-art AI models and the analysis of imaging features, patient characteristics, and anatomical regions on segmentation results, we were able to obtain new insights and provide recommendations for optimized social-technical setting, including clinical data acquisition and the requirements for future development of AI-based segmentation. To begin, our study required the creation of a benchmark dataset, as directly using public datasets or random sampling of private cases would not have been appropriate. Public datasets may overlap with the training data of the models under evaluation, and random sampling of private cases could not ensure a balance of imaging and patient features necessary for statistical analysis. Therefore, we built our benchmark dataset based on real-world scenarios where AI models are applied to end users, and determined the required size with a sample size calculation. Previous studies have shown that resampling could simulate multiple CBCT/CT scans from the same patient in a different image reconstruction settings^27^. Rotational movements of the patient’s head could also be simulated using resampling methods^28^. Hence, we have created a quasi-experiment setting by resampling original CT/CBCT scans and manual screening of patient characteristics. This method provides enough data for the LMM to reveal the underlying factors influencing the performance of AI models.

### Regulations on AI models

Among the 20 models selected for this study, the overall segmentation performance of the commercial models that had received MDR/FDA approval was higher than that of the research models (Fig. 2d). This suggests a positive impact of regulatory policies on the commercial model development and deployment process. However, the costs associated with certifying software as a medical device could be substantial. Regardless of the type of model, monitoring post-deployment performance is a critical step in improving safety as well as the effectiveness of AI models in clinical practice^29^. This is also a key feature of the overall product lifecycle approach used by the FDA^30^. As our study demonstrates, end-users should expect degradation in the performance of current static AI models as a result of changes in imaging protocols or changes in patient populations. One possible solution is dynamic fine-tuning of deployed models. However, the changes in performance as well as risk associated with this continuous learning may cause the product’s metrics to differ from those at the time of initial certification, which would pose a significant regulatory challenge^31^. While regulators are actively developing guidance policies for dynamic tuning models, all approved AI tools have been static up to this date^32,33^. Therefore, the optimization of image acquisition protocols may be a viable alternative solution on static models. Furthermore, the identification of patient characteristics and anatomical regions that cause performance declines could lead to a strategy for intervening, both in the development of AI models as well as in their application.

### Imaging factors and modality

The first questions arise in the optimal reconstruction protocol during the acquisition of medical imaging. Our investigation of one of the most versatile human bones, the mandible, suggests several key areas affecting the quality of AI-based bone segmentation. Elevated sharpness, decreased voxel size, and ensuring standardized patient positioning can all improve AI-based segmentation to a certain degree (Fig. 3). The results are in accordance with findings from traditional segmentation algorithms. Puggelli et al.^34^ reconstructed CT scans of porcine tibiae with different kernels and evaluated the segmentation accuracy compared to laser scanning. The results demonstrated that sharp reconstruction kernel accuracy was higher than that of the soft kernel. The reason for that may be because the bone-soft tissue boundary is better defined in these images. Similarly, another study based on the segmentation results on CBCT scans of an AI model of 11 dry mandibles with different voxel sizes, revealed that larger voxels (0.45 mm) resulted in significant segmentation errors compared to smaller voxels (0.15 mm) (surface scans as reference)^35^. In contrast, Huang et al. concluded when applying one single AI model onto 183 CT scans of 11 patients with different voxel sizes, slice thickness and simulated doses, that there is no need for a strict image resolution^19^. Our comprehensive analysis with 20 models, however, underlined that lower sharpness (increased blurriness) as well as larger voxel size may have a negative impact on segmentation performance. This should be considered in the reconstruction protocols when incorporating AI models.

Another important factor is bone rotation during scanning (in our case the mandible). El Bachaoui et al. collected a total of 20 CBCT scans from 5 fresh cadavers at four different positions^36^. They concluded that the effect of sagittal rotation of the head on segmentation accuracy is clinically negligible (manual segmentation as reference). However, this study investigated a limited range of rotations in the sagittal plane only. In contrast, our study included a wide range of combined rotations in all three reference planes. Our results show that bone rotation in the axial and sagittal planes negatively affects the segmentation results (Fig. 3). This finding is probably due to the underlying distribution of the training data used by AI models. Attention should be paid to the standard positioning of the mandible, especially during CT scanning, as there is more freedom of movement for mandible on supine CT scans that lack chin fixation compared to CBCT. If a proper bone positioning cannot be achieved, post processing into a normalized bone position should be considered.

Regarding the imaging modality, most of the models trained with single modal data (CBCT or CT) were also able to segment scans of the other modality. Only one model, which trained solely on CT data, was unable to do so, as it successfully extracted the skull but was unable to separate the mandible from it. Such results indicate that CBCT and CT are interchangeable in this task, likely due to their similar fundamental imaging principles. Nevertheless, AI segmentation on CBCT demonstrated higher accuracy in descriptive statistics, but the AI model was even better at segmenting the CT data in LMM analysis which took multiple factors into account. The main reason for this may be that the original voxel size of CBCT (0.268 mm in average) is smaller than that of CT (0.442 mm in average), and smaller voxels size leads to better segmentation (Fig. 7). Another reason could be the anisotropy of CT voxels, i.e., slice thickness is generally not equal to in-plane voxel size. In previous studies, this negative effect was predominantly observed in the inter-slice direction, with the main areas affected including the cranial side of the condyle, the inferior border of the mandible, and the alveolar ridge, which is also observed in our study^17^. In contrast, LMM considers voxel size and slice thickness as independent factors, avoiding the interference of voxel morphology on modality. In conclusion, the use of high-resolution CT scans with isotropic voxels may further improve bone segmentation results of AI models.

### Patient-related factors and Regions of Interests

Beside image-related factors, patient-related factors may also affect segmentation accuracy. Our results showed slightly better segmentation performance in males (LMM, β = 0.99%, p < 0.001) (Fig. 4). Yet this difference is marginal, it suggests that the AI models can be readily applied to both sexes. Interestingly, the presence of teeth improved segmentation results (for each additional tooth, LMM, β = 0.38%, p < 0.001). A possible explanation is that teeth act as extra anatomical landmarks for the AI models. Lacking teethless training data could also be a reason. Although restorations and implants are typically the source of artifacts, LMM analysis considered artifacts an individual factor, allowing our study to identify the impact of teeth on segmentation outcomes. However, bone pathology and osteosynthesis materials significantly reduced accuracy.

This result aligns to that from the study of Cui et al. of one single AI model, where evaluated on an external dataset of 407 CBCT scans, missing teeth (DSC, –0.8%), malocclusion (DSC, –0.9%), and metal artifacts (DSC, – 2.0%) negatively affected segmentation results^37^.

The accuracy of mandible segmentation varies in different anatomical regions (Table 3). The condyle exhibits lower accuracy, primarily due to its thin cortical bone and low density of cancellous bone, as well as the surrounding high-density cranial base structures. This results in lower contrast in the condylar region, especially in CBCT images^38^. This was confirmed by our LMM analysis across anatomical regions, where the segmentation performance of the condylar region in CT images is improved by 8.59% in DSC (LMM, β = 8.35%, p < 0.001) compared to CBCT images, while the improvement of the whole mandible segmentation is merely 4.1% (LMM, β = 4.13%, p < 0.001)(Fig. 7). The mandible body also exhibits a higher degree of error in segmentation, which may partially be attributed to the presence of artifacts from the crowns and brackets^39^. Another reason for the drop in the performance on the mandibular body is the discontinuity of the mandible, often accompanied by large osteosynthesis reconstruction plates (Fig. 5 Fig. 6). This could lead to a partial segmentation failure, which in turn severely affects the overall segmentation performance of the mandibular body.

Ideally, AI segmentation models should not be sensitive to reconstruction protocols, patient factors, and anatomical regions, which are highly variable in a socio-technical system. However, due to the limitations in architecture and training data, the current models have not yet reached this goal. Nevertheless, according to our findings, the segmentation performance of the model can be improved by optimizing the imaging protocol. Simulated calculation with results from LMMs suggested that with a recommended protocol (CT scan, sharpness of about 5000 HU/mm, voxel size of 0.5 mm, and neutral bone position), an increase of 9.02% in DSC for AI segmentation can be expected, comparing to the worst combination. In terms of patient characteristics, AI segmentation on a young male with complete dentition, without artifacts, pathology, or osteosynthesis, the DSC would increase by 16.59% compared to the worst combination of features. With these two aspects into account, the difference in DSC between the cases adapted most to fitting predicted requirements of AI models in general and those least adapted would be 25%. A real pair of examples can be found in our dataset (Case 21 and Case 78, Supplementary Table 2), where the mean DSC for AI segmentation of the original volume was 71.82% and 91.49%, respectively, with a difference of 19.67%. This 20% difference in DSC is substantial in terms of workload since cases with DSC above 90% require minor adjustment and those below 75% need intensive manual involvement (Figure 2a).

### Recommendations and Requirements

To narrow this performance gap in clinical practice, collaboration between clinicians and AI developers must focus on mutual adjustments informed by real-world needs. Clinicians can optimize imaging protocols to align with current AI capabilities, while developers should prioritize the requirements that address recurring clinical challenges.

For clinicians, understanding the technical limits of AI models is critical. To improve bone segmentation outcomes, we recommend using CT scans with small, isotropic voxels (0.5 mm or smaller) and high-sharpness protocols when possible. In terms of modality, clinicians should be aware of the potential performance drop in susceptible regions like condyles in CBCT. Also, ensure target bones are positioned neutrally during scans, if not possible (e.g. trauma), adjust the images to a standard orientation before segmentation. In cases with edentulous mandible, large implants, or bone pathologies, clinicians should expect lower accuracy and prepare for manual corrections.

For AI developers, the next-gen models should be stable in performance even when faced with non-ideal clinical conditions. This includes robustness to patient features like bone pathology and osteosynthesis. Considering the sparsity of specific patient group, synthetics data can be a viable option. Segmentation performance in complex anatomical regions (e.g. condyles) should be prioritized, which could be achieved through regionally weighted loss functions or adversarial training for specific structures. In addition, models should explicitly flag uncertain or low-confidence segmentation regions by heatmaps or scores to guide clinician review, particularly in high-risk cases involving bone pathologies or surgical planning.

### Limitation

Our study recruited the largest number of AI models to date and comprehensively analyzed the socio-technical factors including patient factors and imaging factors on segmentation performance. However, one limitation of the study is that we focused on bone segmentation only, which is only one but important fraction of the human anatomy. It would be interesting to see similar investigations into soft tissue segmentation (e.g. hearts, lungs and livers). This may involve analyzing the performance of AI models in various imaging modalities commonly used on soft tissue such as MRI or 3D ultrasound. The impact of factors such as tissue deformation, movement artifacts and inter-patient variability on segmentation results could be factors to be further assessed. In addition, our dataset did not include cases under the age of 18 years because they are not common cases for mandibular bone segmentation. This prevented us from fully capturing anatomical variability in all clinical situations, especially in patients who grow and develop during childhood and adolescence.

### Future work

On our benchmark dataset, the current models still have a certain number of unsatisfying segmentation results, and clinicians need to refine them manually using various tools (Figure 2a). Integrating models with interactive tools (e.g., SAM^40^ and MedSAM^41^) could streamline this “last mile” by allowing clinicians to correct errors via intuitive prompts. This study only briefly investigated the basic architecture used by the models, and due to confidentiality reasons, we were not able to examine in detail the configuration of the training parameters of each model. As a result, the impact of these technical specifications, in addition to the black-box characteristics of AI models, on segmentation accuracy is still not fully understood. Future research should explore these factors, potentially by collaborating to configure models and data in a controlled environment for further experiments.

## Conclusion

This multi-center study shows that the performance of AI mandible segmentation is dynamically shaped by socio-technical factors, including imaging protocols, patient-specific factors and anatomical complexity. Two pillars are essential to the success of clinical translation of AI models: clinicians should adapt their workflows to the current limitations of AI, and developers must tackle the upcoming requirements that address persistent clinical challenges. For clinical teams, this means choosing high-resolution CT protocols when possible, ensuring standardized patient positioning and rechecking AI output in cases involving bone pathology or osteosynthesis. For AI developers, the requirements for the next-gen AI segmentation models are summarized from clinical failures. Models must remain robust to common clinical variabilities like rotation. Models should further improve the accuracy of error-prone anatomical regions (e.g., condyles) and provide intuitive uncertainty feedback to guide clinical reviews. These are not standalone checklists but interconnected obligations—only through this dual commitment can AI progress from a static algorithm and technocentric preoccupation to a trustworthy clinical ally in a socio-technical system.

## Declaration

### Author Contributions

Conceptualization, B.P. and K.X.; Methodology, K.X. and B.P.; software, K.X., M.C. and B.P.; validation, Y.L., A.F. and B.P.; formal analysis, All authors; investigation, K.X., B.P., L.G. and M.C.; resources, B.P., R.R., F.H., M.G., J.S., J.X., E.T., T.PA., M.B., N.S., R.T., G.D., C.W., N.V., P.V., Y.G., Z.X., J.B., A.R., T.F., A.L., R.C., S.V., R.I., S.R., D.M., C.S., T.X., S.B., S.N., O.K., S.Z., M.W., O.S., F.T., H.L., A.C. and T.PO.; data curation, K.X., L.G. and B.P.; writing—original draft preparation, K.X.; writing—review and editing, B.P., F.H., R.R., A.H., S.Z., A.L., and the rest of authors; visualization, K.X. and B.P.; supervision, B.P.; project administration, B.P.; funding acquisition, B.P. All authors have read and agreed to the published version of the manuscript.

### Institutional Review Board Statement

The ethics application of the study was approved by the ethics committee at the RWTH Aachen University (approval number 23-272, 26^th^ October 2023, Prof. Dr. Ralf Hausmann).

### Informed Consent Statement

No informed consent was needed due to the use of anonymized retrospective patient data.

### Data Availability

Due to the model anonymity nature of the study, only the evaluation result with code names of the model is made available in our repository. Benchmarking dataset and the model predictions are available on request from the corresponding author.

### Code Availability

The code for dataset preparation and model evaluation were implemented in Python (Version 3.11.0). The source code and R code for statistical analysis is available on GitHub (https://github.com/OMFSdigital/AI_Mandible_Benchmarking).

## Acknowledgments

We thank the anonymous patients whose CT and CBCT scans formed the basis of this study.

## Supplementary

**Supplementary Table 1.** Demographic and image characteristics of the original scans.

| Patient features | CBCT (n=50) | CT (n=50) | Total (n=100) |
| --- | --- | --- | --- |
| **Age** |  |  |  |
| Mean (SD) | 46.240 (24.090) | 50.700 (20.076) | 48.470 (22.175) |
| Range | 19 - 91 | 22 - 85 | 19 - 91 |
| **Gender** |  |  |  |
| f | 25 (50.0%) | 25 (50.0%) | 50 (50.0%) |
| m | 25 (50.0%) | 25 (50.0%) | 50 (50.0%) |
| **Teeth** |  |  |  |
| Full | 20 (40.0%) | 14 (28.0%) | 34 (34.0%) |
| None | 10 (20.0%) | 12 (24.0%) | 22 (22.0%) |
| Partial | 20 (40.0%) | 24 (48.0%) | 44 (44.0%) |
| **Artifacts** |  |  |  |
| No | 25 (50.0%) | 27 (54.0%) | 52 (52.0%) |
| Yes | 25 (50.0%) | 23 (46.0%) | 48 (48.0%) |
| **Bone Pathology** |  |  |  |
| No | 32 (64.0%) | 28 (56.0%) | 60 (60.0%) |
| Yes | 18 (36.0%) | 22 (44.0%) | 40 (40.0%) |
| **Osteosynthesis** |  |  |  |
| No | 40 (80.0%) | 40 (80.0%) | 80 (80.0%) |
| Yes | 10 (20.0%) | 10 (20.0%) | 20 (20.0%) |
| Imaging features | CBCT (N=50) | CT (N=50) | Total (N=100) |
| **Voxel Size** |  |  |  |
| Mean (SD) | 0.268 (0.019) | 0.442 (0.075) | 0.355 (0.103) |
| Range | 0.250 - 0.287 | 0.289 - 0.662 | 0.250 - 0.662 |
| **Slice Thickness** |  |  |  |
| Mean (SD) | 0.268 (0.019) | 0.706 (0.042) | 0.487 (0.222) |
| Range | 0.250 - 0.287 | 0.700 - 1.000 | 0.250 - 1.000 |
| **Device Name** |  |  |  |
| A | 25 (50.0%) | - | 25 (25.0%) |
| B | 25 (50.0%) | - | 25 (25.0%) |
| C | - | 22 (44.0%) | 22 (22.0%) |
| D | - | 14 (28.0%) | 14 (14.0%) |
| E | - | 14 (28.0%) | 14 (14.0%) |

**Supplementary Table 2.** Sample cases showing the best combination of imaging and patient features verses the worst combination. A decline of 19.67% in DSC was observed. *To avoid identification, age ranges were used. The age difference between the two cases was 44 years.

| Factors | Case 21 | Case 78 | Factor Unit | Beta(%) | Effect(%) |
| --- | --- | --- | --- | --- | --- |
| Gender | F | M | - | 0.99 | 0.99 |
| Age* | 65-70 | 20-25 | 5 years | 0.06 | 0.528 |
| Modality | CBCT | CT | - | 0.38 | 6.08 |
| Teeth Count | 0 | 16 | 1 tooth | 0.21 | 0.21 |
| Artifacts | YES | NO | - | 0.70 | 0.7 |
| Bone Pathology | YES | NO | - | 7.89 | 7.89 |
| Osteosynthesis | YES | NO | - | 4.13 | 4.13 |
| Sharpness | 4010.43 | 5326.17 | 500 HU/mm | 0.50 | 1.32 |
| Voxel Size | 0.29 | 0.45 | 0.10 mm | 0.16 | -0.26 |
| Slice Thickness | 0.29 | 0.70 | 0.10 mm | 0.09 | -0.37 |
| Axial Rotation | 1.14 | -4.47 | 5.00° | 0.51 | -0.34 |
| Coronal Rotation | -2.31 | -0.17 | 5.00° | 0.10 | 0.04 |
| Sagittal Rotation | -13.82 | 12.97 | 5.00° | 0.69 | 0.12 |
| **DSC%_Original** | 71.82 | 91.49 |  |  |  |
| **Real_diff (DSC %)** |  |  |  |  | 19.67 |
| **Model_diff (DSC %)** |  |  |  |  | 21.04 |

**Supplementary Table 3.** Case-wise summary. Attached: Supplementary Table 3-CASE_RANKING.xlsx **Supplementary Table 3**. Average performance of the 20 AI segmentation models on the 100 original cases used in the study as well as their resampled versions for each case. The order of the cases is sorted by segmentation performance (DSC, HD95, MASD, NSD) from best to worst.

**Supplementary Table 4.** Resampling Factors. Attached: Supplementary Table 4-CASES_RESAMPLED_FINAL.xlsx **Supplementary Table 4**. Resampling factors used for all 1000 volumes. The first 100 records are the original volumes. VOZ is the magnification of slice thickness and VXY is the magnification of in-plane voxel size. ROTX, ROTY, and ROTZ correspond to sagittal, coronal, and axial rotations, respectively. The columns SHARNESS and NOISE are measurements of sharpness and noise for that volume. See the online study protocol for more details in resampling.

**Supplementary Figure 1.**
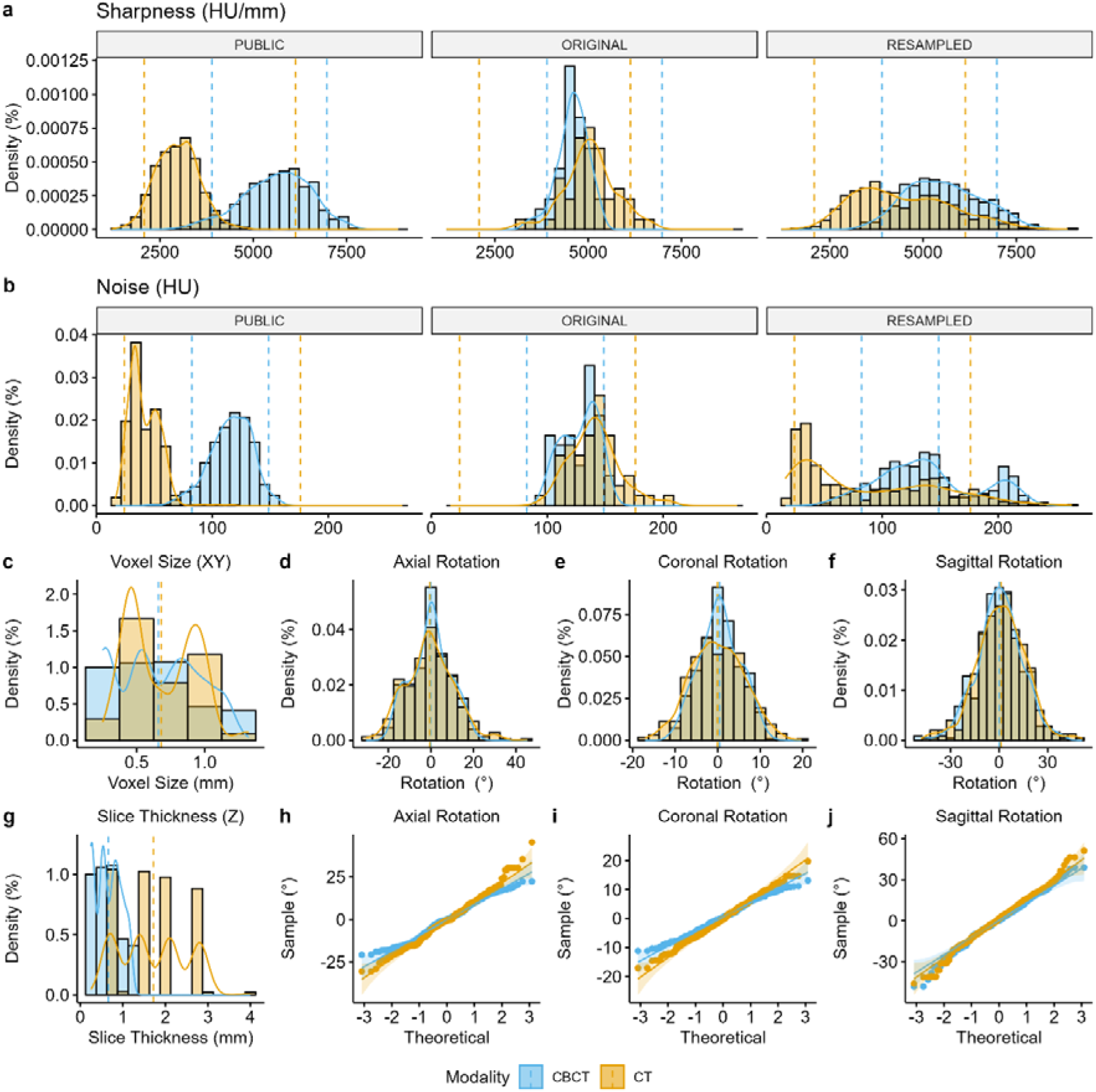
Distribution of imaging features of the final dataset. a,b show the sharpness and noise distributions of the public dataset, the original scans, and the final dataset obtained from resampling, respectively. c and g present the overall voxel size and slice thickness of the final dataset. The final thickness of the CT is not more than 3 mm, and the CBCT voxels remain isotropic after scaling. d-f describe the distribution of the patient’s mandible rotation angles in the final dataset. By adjusting the rotation parameters, the original minus mean value in the sagittal plane due to de-identified cropping have been compensated to approximately zero. h-i are Q-Q plots of the head rotation angle in the three planes, which show that the rotation angle variables are all close to a normal distribution.

**Supplementary Figure 2.**
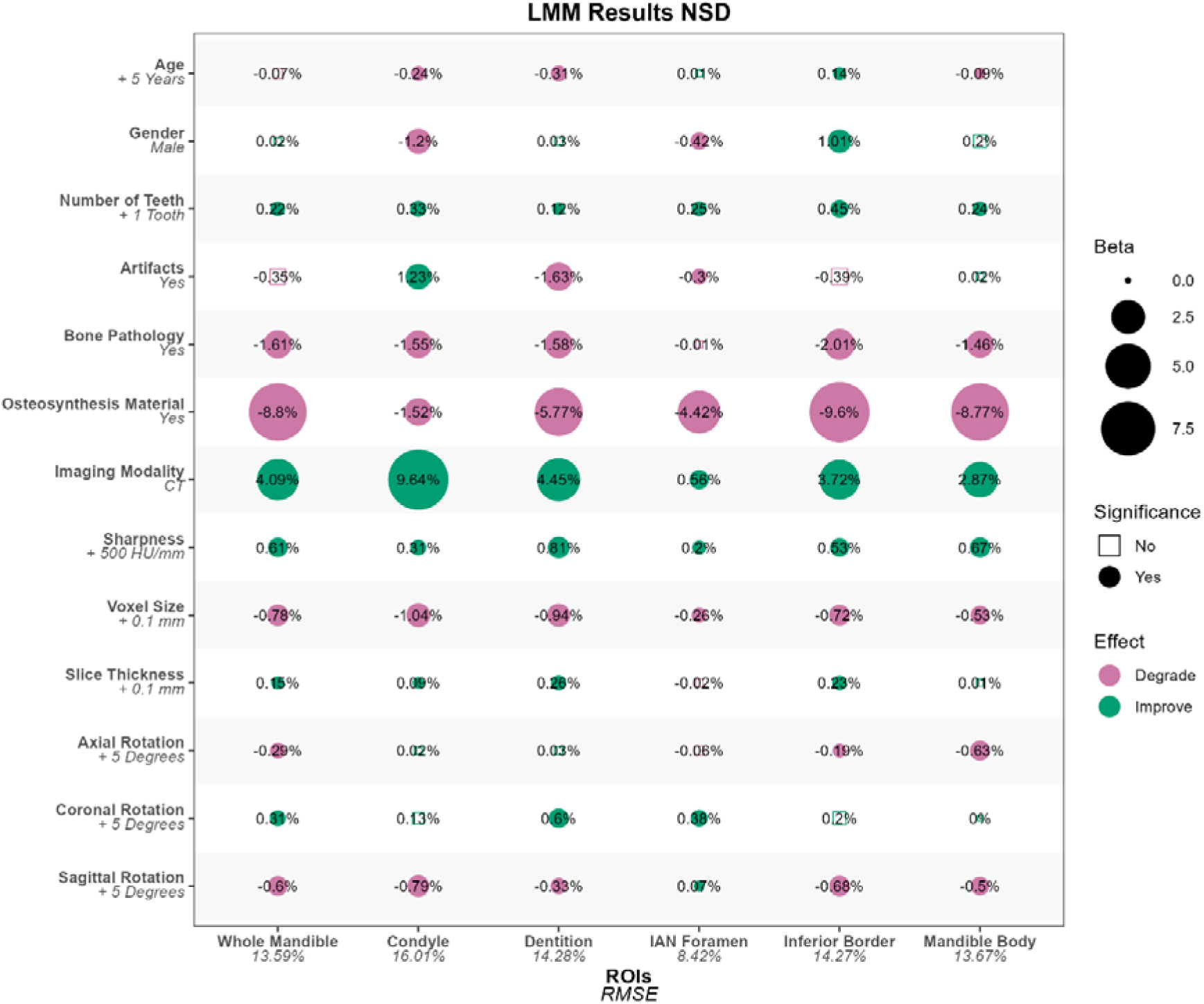
LMMs fitted on evaluation results in NSD% of five ROIs and the whole mandible. Condyles are more affected by modality than in DSC metrics. Factor considered significant when p<0.05.

**Supplementary Figure 3.**
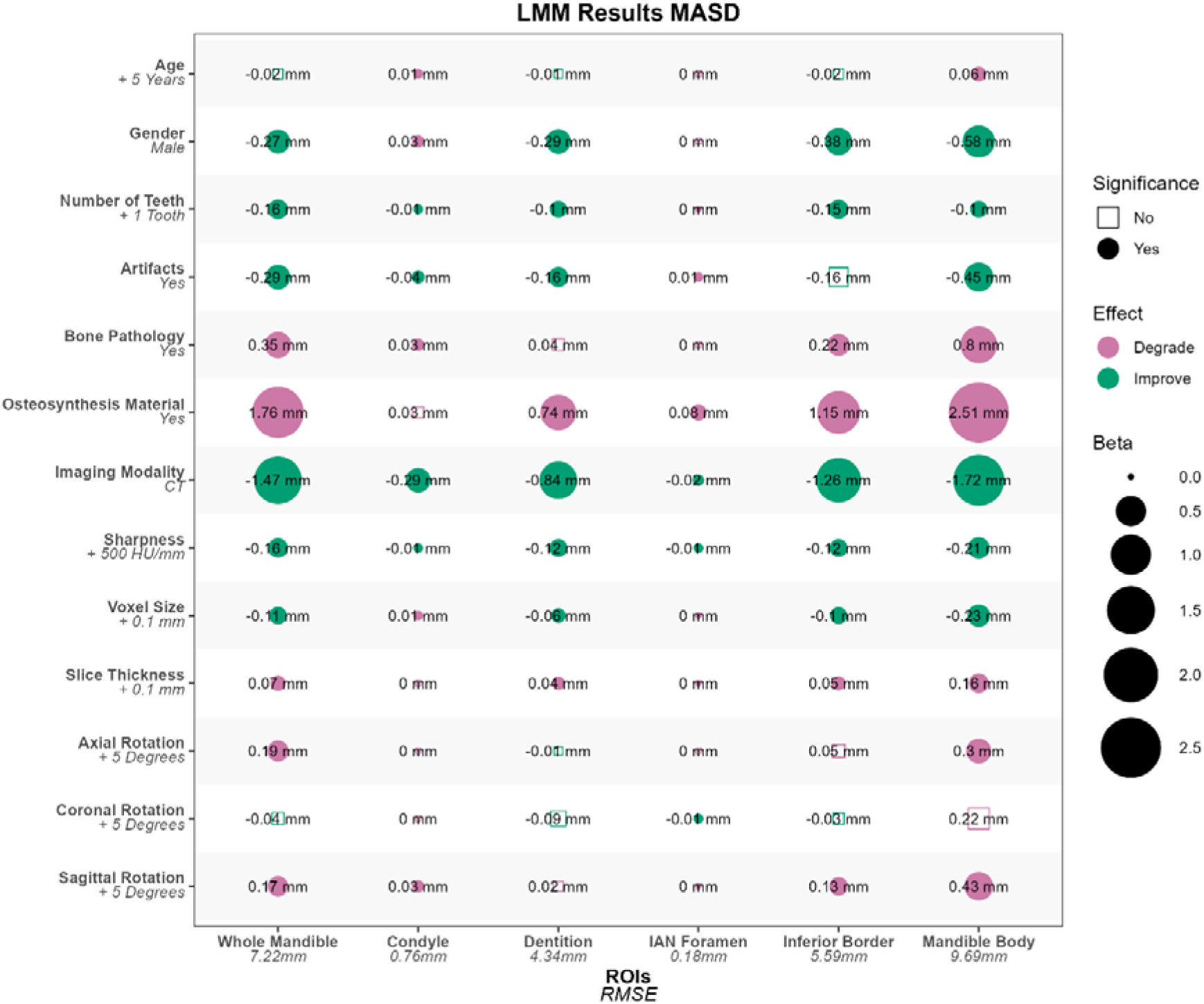
LMMs fitted on evaluation results in MASD (mm) of five ROIs and the whole mandible. Factor considered significant when p<0.05.

**Supplementary Figure 4.**
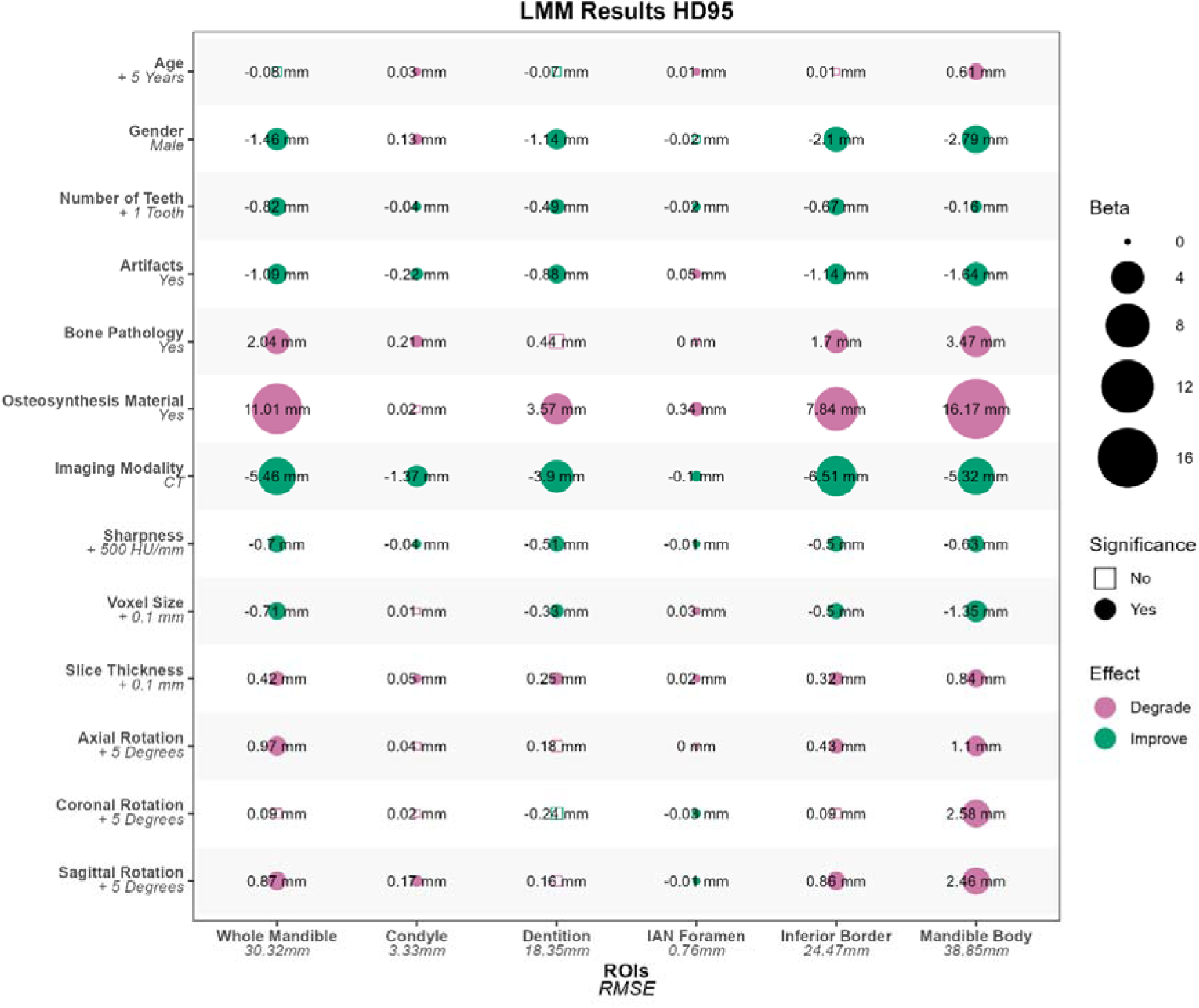
LMMs fitted on evaluation results in HD95 (mm) of five ROIs and the whole mandible. Factor considered significant when p<0.05.

